# Duration and Frequency Mismatch Negativity in Schizophrenia, unaffected first-degree relatives, and healthy controls

**DOI:** 10.1101/2023.05.03.23289437

**Authors:** Anushree Bose, Sri Mahavir Agarwal, Hema Nawani, Venkataram Shivakumar, Vanteemar S. Sreeraj, Janardhanan C. Narayanaswamy, Devvarta Kumar, Ganesan Venkatasubramanian

## Abstract

**Background:** Mismatch negativity (MMN) is elicited upon detecting background irregularities in the sensory environment and subsequent updating of the sensory context. Auditory MMN amplitude is reliably attenuated in schizophrenia patients. However, due to diversity in MMN deviant types (duration, frequency, intensity, gap, etc.), considerable variability exists in MMN findings reported from the early course and chronic samples. MMN is sometimes reported to be impaired or associated with schizotypy, but MMN and schizotypy are yet to be well examined in unaffected first-degree relatives of schizophrenia patients.

**Methods:** Fifty-two schizophrenia patients (SZ) were compared with thirty-six unaffected first-degree relatives (FDR) of schizophrenia patients and thirty-two age and sex-matched healthy controls (HC) on MMN indices using a two-tone passive auditory oddball paradigm with two conditions – duration deviant (MMNd) and frequency deviant (MMNf) event-related potential experiment. SZ sample was further split into two sub-groups 1) early-course/drug-naïve or drug-free (dSZ), and 2) chronic/medicated (cSZ) to examine the effect of illness chronicity and medication on MMN indices. We also checked whether schizotypy scores associated with MMNd and MMNf amplitudes in the FDR group.

**Results:** At baseline, SZ group had significantly diminished MMNd amplitude compared to both HC and FDR groups (p<0.001). The SZ group also had significantly lower MMNd latency than the FDR group (p<0.014). The cSZ and dSZ groups did not differ from each other on MMN amplitude or latency, though cSZ group had lower MMN amplitude. Only cSZ patients showed negative correlation of MMNd amplitude with hallucinations scores and total duration of illness. In FDRs, MMNd and MMNf amplitudes showed negative correlation with the cognitive-perceptual factor of schizotypy.

**Discussion:** Deficient MMNd in SZ patients adds further support to the prediction error estimation abnormalities in schizophrenia. MMNd is a more robust measure than MMNf in differentiating SZ from FDR and HC. MMNd amplitude could be more impaired in hallucinating SZ patients and associate with illness chronicity. Though unaffected FDRs have MMN comparable to healthy controls, higher schizotypy in FDR is associated with lower MMN amplitude. MMN and schizotypy are potentially linked and deserve a nuanced examination.

## INTRODUCTION

Mismatch negativity (MMN) is an electrophysiological index of deviance-detection elicited pre-attentively whenever established contextual regularity in the sensory environment is violated (Näätänen et al., 2001). Auditory MMN is the product of a pre-attentive phenomena that detects background irregularities in the auditory environment. It is elicited from the deviation effect that manifests when immediate auditory input is compared with a memory trace of a previous sound preceding it. As an index of discrimination accuracy, it also reflects the integrity of central auditory system plasticity (Näätänen, 2008). With schizophrenia being empirically investigated as a disorder of neuroplasticity with abnormalities in long-term potentiation changes (McCullumsmith, 2015; McCullumsmith et al., 2004), MMN becomes an intriguing tool for examining the overall integrity of the auditory processing system in Schizophrenia.

MMN deficits across varying stimulus parameters (like pitch/frequency, duration, intensity, and gap) (Avissar et al., 2018; Umbricht and Krljes, 2005) have been consistently reported among SZ patients (Rissling et al., 2010). However, the most studied MMN deviant types are: duration (MMNd) and frequency/pitch (MMNf). Both MMNd (Coffman et al., 2017; Haigh et al., 2017a; Haigh et al., 2017b; Umbricht and Krljes, 2005) and MMNf (Coffman et al., 2017; Giordano et al., 2021; Haigh et al., 2016; Haigh et al., 2017b) amplitudes are widely reported to be suppressed in SZ, however the effect size for deficient MMNd amplitude is most robust (Erickson et al., 2016; Umbricht and Krljes, 2005). Deficient MMN amplitudes are considered to be specific to the genetic expression of SZ (Umbricht et al., 2003). The attenuation in MMN amplitude in SZ patients has been a pervasive observation irrespective of medication status, and chronicity of illness, and is detectable even in the prodromal state (Bodatsch et al., 2011). Research findings indicate that MMN deficits in SZ – (i) are indicative of dysfunction in pre-attentive auditory pre-processing and dysregulated responsiveness of auditory cortices to resource allocation for incoming information (Damaso et al., 2015), (ii) exist irrespective of hallucinatory status (Fisher et al., 2011) and independent of illness duration, psychopathology and cognitive impairments (Giordano et al., 2021), (iii) are present early on (Rudolph et al., 2015), (iv) precede the onset of the disorder itself (Donaldson et al., 2022; Murphy et al., 2020; Perez et al., 2014), (v) are fairly stable for up to five years and foretell poor prognosis, that is, worsening of illness severity, especially auditory verbal hallucinations, and everyday functioning (Donaldson et al., 2022; Giordano et al., 2021; Murphy et al., 2020). MMN indices possess predictive value in gauging psychosis risk (Hamilton et al., 2022; Perez et al., 2014) and determining the transition to psychosis from a prodromal state (Hamilton et al., 2022; Näätänen et al., 2015). Interestingly in several studies, deficient MMN indices (like amplitude and latency) have been found to associate with positive symptoms of SZ (Kärgel et al., 2014) particularly state and trait measures of auditory verbal hallucinations (AVH) (Fisher et al., 2014).

MMN is often examined in the first-degree relatives of SZ patients to study whether MMN is deficient in them or if it predicts a transition to psychosis. Some studies report MMN amplitude to be attenuated in FDR (Jessen et al., 2001; Michie et al., 2002; Şevik et al., 2011) while others report MMN amplitude in FDR is comparable to healthy controls (Ahveninen et al., 2006; Bramon et al., 2004; Donaldson et al., 2021; Hong et al., 2012; Magno et al., 2008). Despite the contradiction, such approaches are essential to understand whether MMN indices can detect schizophrenia-like traits that exist as a continuum in the general population that facilitates early detection and early intervention and alters the disease course.

Trait-level expression of schizophrenia is perhaps best captured in a non-clinical population with schizotypy (Ettinger et al., 2014). Schizotypy is expressed as a cluster of discernible personality traits like unusual perception, suspiciousness, odd beliefs, eccentric behaviour, magical thinking, social isolation, and constricted affect which broadly relate to positive, negative and disorganized symptom categories of schizophrenia (Raine and Benishay, 1995). As MMN is deemed to be a trait marker for psychosis, examining unaffected first-degree relatives of SZ patients can shed light on whether MMN is a sensitive enough index to facilitate early intervention. MMN deficits have been noted or examined in individuals with schizotypal personality disorder (Niznikiewicz et al., 2009), siblings of SZ patients with schizotypy (Donaldson et al., 2021) and healthy controls expressing schizotypy (Broyd et al., 2016). However, whether and how MMN and schizotypy are related is yet to be properly investigated.

In this study, we examined MMN indices in SZ patients in comparison to healthy controls (HC) and unaffected first-degree relatives (FDR). We hypothesized SZ patients to have worse MMN amplitude and latency compared to the FDR and HC. We also expected the unaffected FDR to show marginal to no deficit in MMN in comparison to HC. We split the SZ sample into two sub-groups 1) early-course/drug-naïve or drug-free – dSZ group, and 2) chronic/medicated – cSZ group to examine the effect of illness chronicity and medication on MMN indices. We expected clinical symptoms to show a negative correlation with MMN indices in SZ such that higher psychopathology or illness chronicity will associate with lower MMN amplitude, especially for the chronic/medicated patients, that, is cSZ group. We also examined schizotypal personality traits in the unaffected first-degree relatives and hypothesized that participants with higher schizotypal traits will have lower MMN amplitudes.

## METHODS

### Participants

Fifty-two schizophrenia patients, attending the clinical services of the National Institute of Mental Health And Neuro Sciences (NIMHANS), Bengaluru, participated in this study. Trained interviewers ascertained diagnosis of schizophrenia as per DSM–IV TR in patient participants using Mini International Neuropsychiatric Interview (Sheehan et al., 1998) Experienced psychiatrist (GVS/JCN) independently re-affirmed the diagnosis for each patient. Thirty-six unaffected first-degree relatives of schizophrenia patients also consented to take part in this study. Out of these thirty-six unaffected FDR, schizotypy scores of thirty-three participants were previously reported in a neuroimaging study (Kalmady et al., 2020). Thirty-two healthy participants were recruited as a control group. Except for two participants, one HC and one FDR who were ambidextrous, the remainder of the participants were right-handed. Edinburgh Handedness Inventory (Oldfield, 1971) was used to confirm the handedness of each participant. The institute ethics committee had reviewed and approved this study. Written informed consent was received from every participant before taking part in this study.

### Clinical Assessment

Detailed clinical history was obtained for the SZ patients. Comprehensive assessment of the positive and negative symptom psychopathology in SZ patients was done with Scale for Assessment of Positive Symptoms (SAPS) (Andreasen, 1984) and Scale for assessment of Negative Symptoms (SANS) (Andreasen, 1989). The hallucination sub-score of SAPS was separately taken and used in statistical analysis to explore the potential link between MMN indices and hallucination. Medication dosage was noted and olanzapine equivalent was calculated as per prescribed guidelines (Leucht et al., 2016). Assessment of schizotypal traits in unaffected FDR was done with Schizotypal Personality Questionnaire-Brief (SPQ-B) (Raine and Benishay, 1995). For unplanned post-hoc analyses, we calculated the sub-scores for three dimensions/factors of SPQ-B scale by adding up corresponding item scores: cognitive-perceptual (item no. 2, 4, 5, 9, 10, 12, 16, and 17), interpersonal (item no. 1, 7, 11, 14, 15, 18, 21 and 22) and disorganized (item no. 3, 6, 8, 13, 19 and 20) (Raine and Benishay, 1995).

### Assessment of Mismatch Negativity (MMN)

DATA ACQUISITION: Event-related potential based assessment of auditory MMN was done as per previous descriptions (Chen et al., 2014) in a dark, electrically shielded room. Paradigm was designed with and administered through STIM 2.0 stimulation presentation and experimental design system (Neuroscan, Compumedics). Auditory stimuli were delivered through disposable soft-tip in-ear headphones while subjects watched a nature-themed muted video. Instruction to the participants were: concentrate on the video and ignore the sounds coming through the earphones. They were asked simple questions related to the video at the end of the experiment to check whether they had paid attention to the video. To ensure clear deliverance of auditory stimuli, the sound intensity was set at 70 dB which was adequately above the auditory threshold of all participants. The MMN task had two conditions – Duration deviant, MMNd (1000 Hz, 150 msec) and Frequency deviant, MMNf (1100 Hz, 50 msec). The standard tone in both conditions was of 1000 Hz, 50 msec. Each condition had a total of 750 trials, with a standard to deviant ratio of 9:1, an inter-stimulus interval of 500 msec, with pseudo-random order of stimuli presentation. Each block was of approximately 7 minutes. One block of both conditions was administered consecutively with a gap of about two minutes in between for sensory wash-out. The order of presentation of MMNd and MMNf blocks was counterbalanced across subjects. Continuous EEG data was recorded at a sampling rate of 1000 Hz; bandpass filtered at 0.01-300 Hz. Data was recorded in Curry 7.05 software (Neuroscan, Compumedics) from central electrodes (Fz, FCz, Cz, CPz, Pz, Oz), ground electrode (placed on forehead), double mastoids (used as reference), two bipolar channels VEOG (vertical electrooculogram) and HEOG (horizontal electrooculogram).

DATA PROCESSING: Off-line signal processing was done using BrainVision Analyzer 2.2.2 (Brain Products, GmbH) after converting EEG data files from Curry format to BrainVision format using EEGLab and relevant plug-ins (loadcurry v3.3.0 and bva-io v1.71). Data was band pass filtered at 1 to 30 Hz. Gratton & Coles method (Gratton et al., 1983) was applied to correct the eye blink activity by using signals from VEOG & HEOG. The time segments with movement or other artefacts were excluded from further analysis using a semi-automated (interactive view) artifact rejection algorithm whereby trials were rejected when (a) the absolute voltage difference between two data points exceeded ±100 μV, (b) the difference between the maximum and minimum value in a 200ms segment exceeded ±100 μV, (c) the amplitude exceeded ±100 μV, (d) the difference between the maximum and minimum in an interval of 100 ms exceeded 0.5 μV and (e) deemed noisy upon visual inspection. Artifact-free, continuous EEG data was segmented into fixed time segments or epochs of 550 msec (from – 50 msec to 500 msec relative to the stimuli onset). The pre-stimulus interval was used for baseline correction. These epochs were averaged separately for both standard and deviant stimuli. MMN waveform was generated by subtracting the ERP response to standard’s average from the ERP response to deviant’s average. Automated detection for peak amplitude and its latency was done within the window length of 150 to 250 msec.

### Statistical Analysis

Statistical tests were performed with SPSS 28 (https://www.ibm.com/). After subjecting demographic, clinical and neurophysiological variables to normality test and outlier detection, parametric (One–way ANOVA, independent sample t–test, etc.) or equivalent non–parametric tests (Kruskal–Wallis test for independent samples, Mann–Whitney U test, etc.) were performed to test for statistically relevant groups differences. With ANOVA, Games–Howell Post–Hoc test was performed to explore the pairwise between-group differences as the groups had unequal variances and unequal sample sizes. Alongside independent sample Kruskal Wallis test, Dunn–Bonferroni post hoc tests was performed to test for between group differences as the subsets for pair-wise comparisons were small and had unequal variances. For bivariate correlational analysis, we chose a one-tailed test to test our directional hypothesis. If both the variables were normally distributed, we reported Pearson’s r; if not, we reported Spearman’s rho. The exploratory analyses were not corrected for multiple comparisons.

## RESULTS

### Baseline Comparison of SZ with FDR and HC

Demographics details for SZ, FDR and HC and clinical details for SZ are provided in Table 1. None of the observations from any of the variables—demographic or clinical or neurophysiological—were found to be a noteworthy outlier warranting exclusion from analyses. Disparity in age (F=4.26; p=0.016) and education [*χ*^**2**^(2)=22.50; p<0.001] were noted among the groups. SZ participants were significantly older than HC (i-j=4.031, p=0.02), however SZ group was comparable to FDR group, and FDR group was comparable to HC group in age. HC group had significantly higher education than both the SZ (i-j=–34.50; p<0.001) and FDR (i-j=–32.86; p<0.001) groups.

**Table 1:**
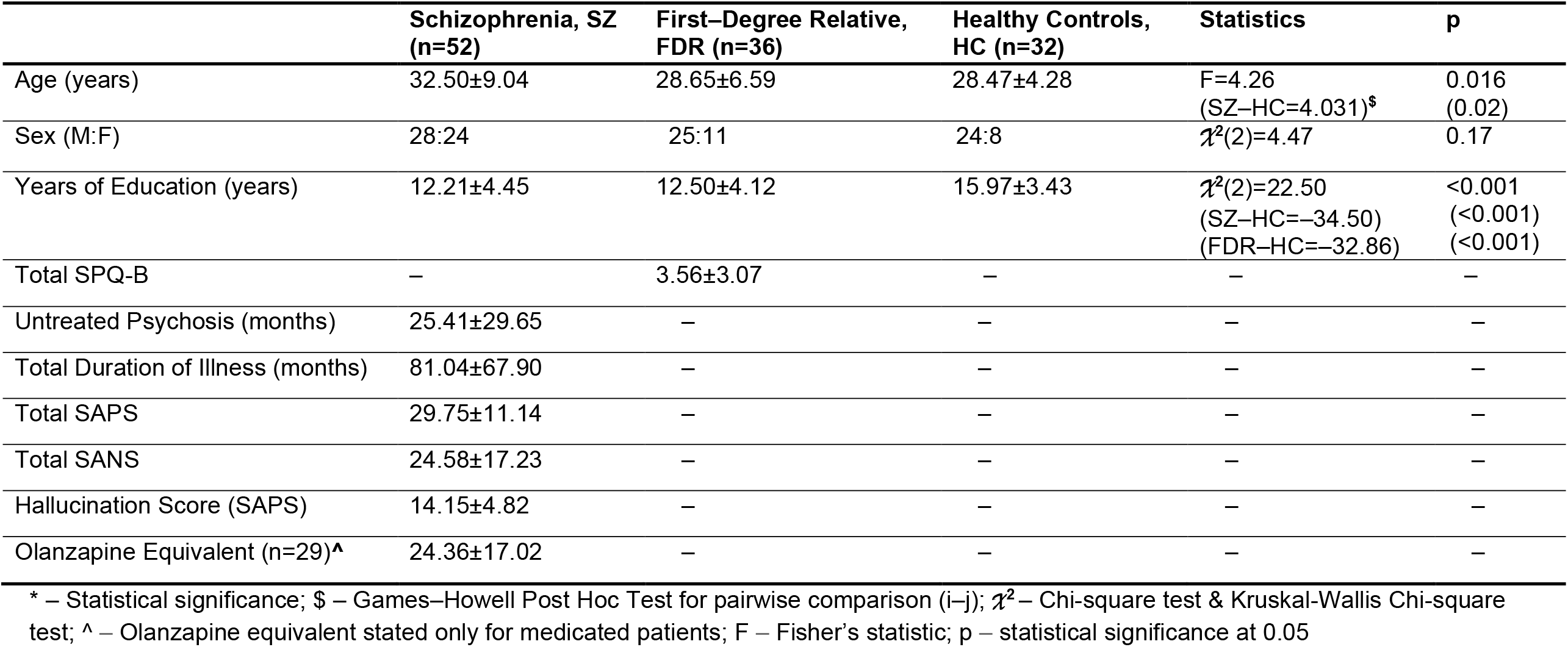
Demographics and clinical variables for SZ, FDR and HC.

|  | Schizophrenia, SZ<br>(n=52) | First-Degree Relative,<br>FDR (n=36) | Healthy Controls,<br>HC (n=32) | Statistics | p |
| --- | --- | --- | --- | --- | --- |
| Age (years) | 32.50±9.04 | 28.65±6.59 | 28.47±4.28 | F=4.26<br>(SZ–HC=4.031)<sup>\$</sup> | 0.016<br>(0.02) |
| Sex (M:F) | 28:24 | 25:11 | 24:8 | $\chi^2(2)=4.47$ | 0.17 |
| Years of Education (years) | 12.21±4.45 | 12.50±4.12 | 15.97±3.43 | $\chi^2(2)=22.50$<br>(SZ–HC=–34.50)<br>(FDR–HC=–32.86) | <0.001<br>(<0.001)<br>(<0.001) |
| Total SPQ-B | – | 3.56±3.07 | – | – | – |
| Untreated Psychosis (months) | 25.41±29.65 | – | – | – | – |
| Total Duration of Illness (months) | 81.04±67.90 | – | – | – | – |
| Total SAPS | 29.75±11.14 | – | – | – | – |
| Total SANS | 24.58±17.23 | – | – | – | – |
| Hallucination Score (SAPS) | 14.15±4.82 | – | – | – | – |
| Olanzapine Equivalent (n=29)<sup>^</sup> | 24.36±17.02 | – | – | – | – |

\* – Statistical significance; \$ – Games–Howell Post Hoc Test for pairwise comparison (i–j); $\chi^2$ – Chi-square test & Kruskal-Wallis Chi-square test; ^ – Olanzapine equivalent stated only for medicated patients; F – Fisher’s statistic; p – statistical significance at 0.05

Out of the four MMN indices, only MMNf amplitude was found to be normally distributed across the three groups (SZ, FDR and HC), so one–way ANOVA with Games–Howell Post–Hoc test was performed. For other MMN variables, that are—MMNd amplitude, MMNd latency and MMNf latency— independent sample Kruskal Wallis with Dunn–Bonferroni post hoc tests were done. Bonferroni corrected p level was set at 0.01 (0.05/4).

MMNd amplitude significantly varied across the groups [*χ*^**2**^(2)=21.41;p<0.001] (Table 2; Figure 1a). SZ participants had significantly diminished MMNd amplitude compared to both HC (mean difference=30.17; p<0.001) and FDR (mean difference=29.17; p<0.001) groups. However, the mean MMNd amplitude between HC and FDR groups were comparable. The mean MMNd latency for SZ groups was much lower than the FDR group (mean difference=34.50; p<0.014). No between–group differences were noted for MMNf amplitude and MMNf latency (Table 2; Figure 1b). For the SZ group, none of the clinical variables showed significant correlation with any of the MMN-related variables.

**Table 2:** MMN variables for Schizophrenia (SZ; n=52), unaffected first-degree relatives (FDR; n=36) and Healthy controls (HC; n=32)

|  | SZ (M±SD) | FDR (M±SD) | HC (M±SD) | Statistics | p |
| --- | --- | --- | --- | --- | --- |
| MMNd Amplitude (μV) | -1.80±1.22 | -3.06±1.49 | -2.95±1.18 | $\chi^2(2)=21.41$<br>(SZ-HC= -30.17)<sup>#</sup><br>(SZ-FDR= -29.17)<sup>#</sup> | <0.001*<br>(<0.001)<br>(<0.001) |
| MMNd Latency (msec) | 187.75±30.14 | 202.58±24.37 | 193.37±22.88 | $\chi^2(2)=8.03$<br>(SZ-FDR= -34.50)<sup>#</sup> | 0.018*<br>(0.014) |
| MMNf Amplitude (μV) | -1.59±1.12 | -1.86±1.09 | -1.46±1.13 | F=1.19 | 0.30 |
| MMNf Latency (msec) | 205.56±33.52 | 191.69±27.09 | 192.94±31.62 | $\chi^2(2)=6.06$ | 0.048 |

**Figure 1:**
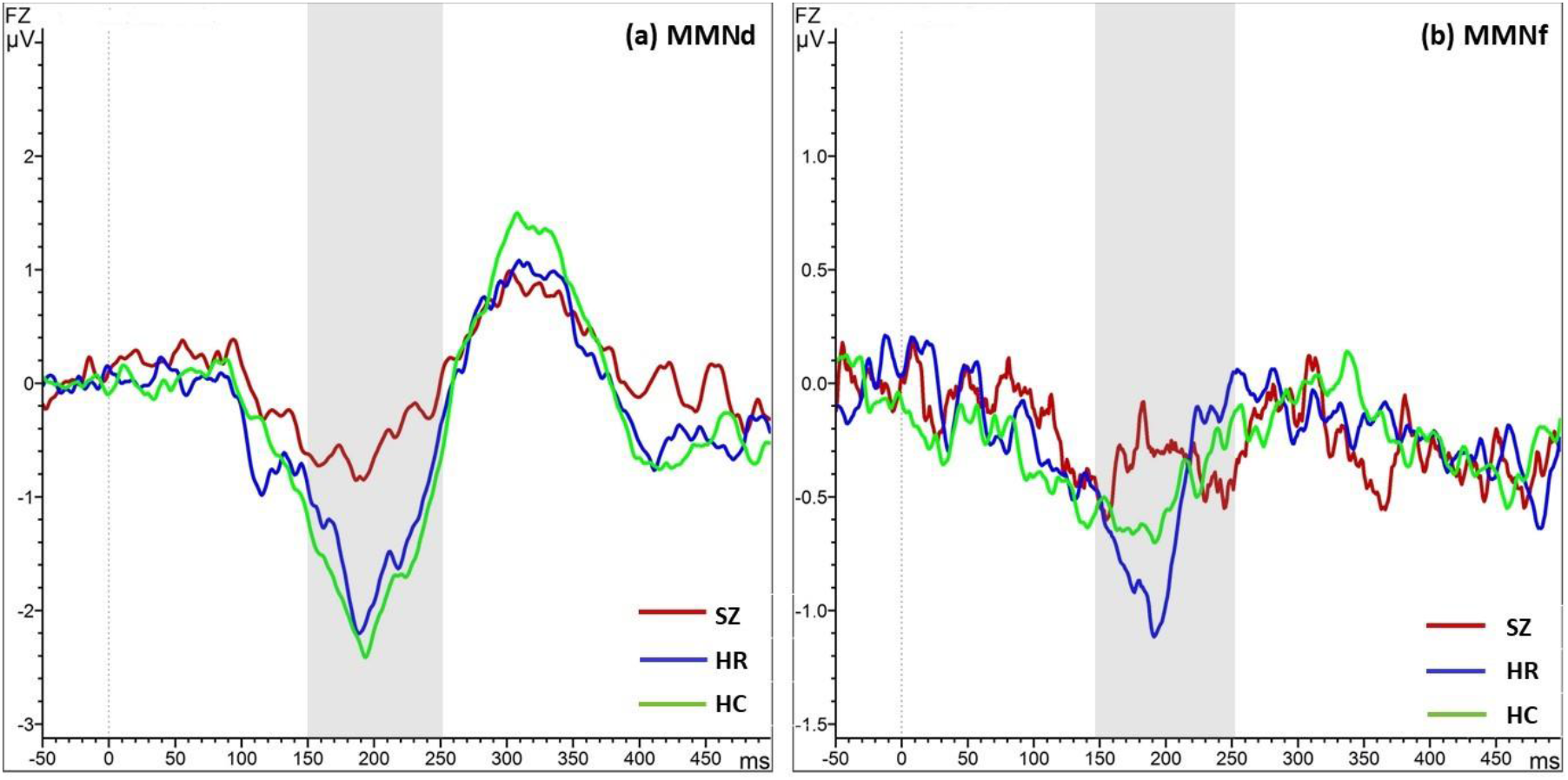
Grand averaged (a) MMNd and (b) MMNf waveforms for SZ, HR and HC. The scale/amplitude gain for MMNd is from +3 μV tp -3 μV, and for MMNf, the scale/amplitude gain is from +2μV to -1.5 μV

Within the SZ group, we recognized a subset of early course/drug–naïve or drug–free patients (dSZ; n=23) and chronic/medicated patients (cSZ; n=29). Exploratory analyses were done to investigate whether medication status and illness chronicity affected any of the MMN indices of interest (Supplementary Table 1). The Kruskal Wallis test for four independent samples, namely: early-course/drug–naïve or drug–free SZ (dSZ), chronic/medicated SZ (cSZ), FDR and HC yielded similar results. Dunn’s Bonferroni–corrected Post–Hoc Test for pairwise comparison did not indicate any significant group differences in any of the ERP variables between the dSZ and cSZ groups. However, the post–hoc pairwise comparisons indicate that the magnitude of mean group differences was higher for cSZ–HC (i-j=–32.77, p=0.001) and cSZ–FDR (i-j=–31.77, p=0.001) than dSZ–HC (i-j=–26.62, p=0.034) and dSZ–FDR (i-j=–25.62, p=0.039) for MMNd amplitude.

Illness chronicity (U=143; p<0.001) and duration of untreated psychosis (U=164; p=0.002) were significantly higher for the cSZ group than dSZ group. The cSZ group had significantly higher positive symptom psychopathology than the dSZ group as indicated by the baseline Hallucination sub–score (t=4.07; p<0.001) and SAPS (t=3.19; p=0.002) scores. Mean SANS scores at baseline did not significantly differ between the two groups (t=0.50; p=0.95). For the cSZ group, significant negative correlation for MMNd amplitude with 1) severity of hallucinations (Hallucination sub-score, SAPS) (r=– 0.41; uncorr p=0.014; one-tailed) and 2) total duration of illness (r=–0.35; uncorr p=0.032; one-tailed). None of the other clinical variables showed correlation with MMN indices for the cSZ or dSZ groups. Medication dosage did not correlate with any of the clinical or neurophysiological variables.

In the unaffected FDR group, mean SPQ-B score was 3.56 (SD=3.07). The SPQ-B total score showed a negative correlation with MMNf amplitude (spearman rank coefficient, ρ=0.28; uncorr p=0.047; one-tailed). Intrigued, we looked whether specific factors of SPQ-B scale explained the correlation. The cognitive-perceptual factor of SPQ-B showed negative correlation with both MMNf (spearman rank coefficient, ρ=0.30; uncorr p=0.037; one-tailed) and MMNd amplitudes (spearman rank coefficient, ρ=0.30; uncorr p=0.037; one-tailed). The interpersonal factor of SPQ-B showed negative correlation with MMNf amplitude (spearman rank coefficient, ρ=0.31; uncorr p=0.032; one-tailed). The disorganized factor of SPQ-B did not show any such correlation with the MMN indices.

## DISCUSSION

Robust MMN deficits are consistently reported in both early-course and chronic SZ patients. Despite the increasing variability and complexity in MMN paradigms, suppressed duration deviant and frequency deviant MMN amplitudes in SZ in comparison to healthy controls remain the most widely reported findings in literature. However, MMN findings from sub-clinical populations like unaffected FDR is not so unequivocal. Furthermore, a few reports have noted MMN deficits in individuals with schizotypy as well. Our goal was to explore MMNd and MMNf deficits cohesively in a sample of SZ patients (drug naïve/drug free and medicated/chronic) in comparison to HC and unaffected FDR. We also explored whether features of schizotypy correlate with MMN amplitudes in unaffected FDR.

Our primary hypothesis was SZ patients will show suppressed MMNd and MMNf amplitudes in comparison to HC and unaffected FDR. As anticipated, SZ patients had attenuated MMNd amplitude in comparison to both the FDR and HC groups but the SZ group had lower MMNd latency compared to the FDR group. Among the three groups, SZ patients had both the lowest mean amplitude and mean latency for MMNd while FDR group had the highest means for MMNd amplitude and latency. Attenuated mean amplitude for MMNd in SZ has been well documented by previous studies and metanalyses (Coffman et al., 2017; Haigh et al., 2017a; Haigh et al., 2017b; Umbricht and Krljes, 2005). Compared to HC, the MMNd deficit in SZ is reportedly both more pronounced and stable than MMNf deficit (Kathmann et al., 1999; Umbricht and Krljes, 2005). With respect to FDR, our observations reiterate previous findings. An earlier study had reported findings similar to ours, that is, MMNd amplitude in SZ patients is attenuated in comparison to both FDR and HC but MMNd amplitude did not differ between the FDR and HC (Bramon et al., 2004). Like us, some studies have noted unimpaired MMNd amplitude in unaffected FDR (Bramon et al., 2004; Donaldson et al., 2021; Magno et al., 2008). Literature also indicates symptomatic clinical-high risk samples, especially those who later transition to psychosis, to have MMN amplitudes that are intermediate between SZ and HC (Bodatsch et al., 2011; Brockhaus-Dumke et al., 2005; Shaikh et al., 2012); our FDR sample was screened for psychopathology, had low schizotypy score and could not be followed-up long enough to note their transition to psychosis status, and hence do not fall within these categories. However, interestingly, previous studies note that non-transitioning high-risk for psychosis samples did not show deficient MMNf or MMNd in comparison to HC (Bodatsch et al., 2011; Sehatpour et al., 2020).

Contrary to our hypothesis, neither the mean amplitude nor the mean latency for MMNf was found to be significantly different among the groups. There are a few studies that have reported pitch/frequency deviant MMN amplitude to be smaller in SZ compared to HC. However, these studies had a higher frequency difference between the standard and the deviant tone (**Δ**200Hz) compared to us (**Δ**100Hz) (Coffman et al., 2017; Giordano et al., 2021; Haigh et al., 2016; Haigh et al., 2017b). Negative reports like ours also exist (Magno et al., 2008) especially for recent-onset/first-episode psychosis patients (Chen et al., 2014; Haigh et al., 2017a; Salisbury et al., 2017; Salisbury et al., 2002; Weigl et al., 2016; Xiong et al., 2019). Yet, even after we split the SZ group into chronic/medicated (cSZ) and early-course/drug-naïve or drug-free (dSZ) sub-groups, we did not detect reduction in MMNf amplitude in cSZ or dSZ in comparison to HC and FDR groups. As per metanalyses, in SZ, the effect size for MMNd deficit is significantly higher than the effect size of MMNf deficit (Erickson et al., 2016; Umbricht and Krljes, 2005). Also, MMNf has been noted to be impaired in a sub-type of SZ patients with poor functional outcome who are partially or completely dependent on assisted care (Lee et al., 2018). Though we did not systematically measure functional outcome in our sample, our SZ sample was clinically stable and most of the SZ participants were either outpatients or were admitted because they were out stationed. Taken together, the prevailing literature on weaker potency of MMNf and our sample’s characteristics seem to explain why MMNf amplitude did not differ between SZ, HC and FDR in this study.

The cSZ and dSZ groups did not show statistically significant difference from each other on MMNd indices but both showed attenuated MMNd amplitude in comparison to HC and FDR as previous studies have noted for early course/first-episode psychosis patients (Murphy et al., 2020; Xiong et al., 2019). The cSZ had more attenuated MMNd amplitude than dSZ. This finding is similar to a recent report where SZ patients were divided into four sub-groups based on illness chronicity and MMNd amplitude did not differ among these SZ groups but all of these SZ groups had attenuated MMN amplitudes in comparison to HC, and the most chronic SZ group had the lowest MMN amplitude (Giordano et al., 2021). Like us, several studies (Magno et al., 2008; Murphy et al., 2020; Xiong et al., 2019) and one meta-analysis (Erickson et al., 2016) have noted chronic SZ patients to have more deficient mean MMNd amplitude than FES (first episode psychosis). Also, a recent report indicates that 1^st^ generation antipsychotics or a combination of 1^st^ and 2^nd^ generation antipsychotics significantly lower MMNd amplitude in medicated SZ patients in comparison to unmedicated SZ patients (Light et al., 2015). Thus, it is reasonable to assume that factors like disease course and medication-type effects may have contributed to the lack of significant difference between MMNd amplitude in dSZ and cSZ.

Among the SZ sub-groups, only the cSZ group showed negative correlation between hallucination scores and MMNd amplitude. This finding is supported by a recent trans-diagnostic study that reported severe hallucinations (in samples across the psychosis spectrum) correlate with decreased MMNd (Donaldson et al., 2020). Other MMN studies reporting an association between auditory hallucination and MMN (gap deviant) amplitude had used a complex multi-feature MMN paradigm; thus drawing a parallel is not ideal (Fisher et al., 2011; Fisher et al., 2012).

Interestingly, cSZ showed a negative correlation between MMNd amplitude and the total duration of illness but not the dSZ group. The duration of untreated psychosis did not correlate with any MMN measures. Given the postulated role of MMN as an indicator of predictive coding aberrations in SZ (Fong et al., 2020; Kirihara et al., 2020), this suggests that MMNd not just worsens over the course of illness but it may be reflective of progressive worsening of prediction error in response to persistence of illness. Existing evidence points that in early course SZ patients, MMN deficits are not associated with illness chronicity or medication. Thus, present findings add to the increasing pool of evidence for SZ pathogenesis to involve aberrations in predictive processing (Adams et al., 2013; Friston, 2023; Sterzer et al., 2018).

None of the MMNd or MMNf indices in SZ showed a significant correlation with any of the demographic variables or positive/negative symptom scores. Though robust metanalyses (Erickson et al., 2017; Erickson et al., 2016) support a lack of association between SZ symptoms, illness chronicity and MMN indices; very few studies have reported negative correlation between MMN indices and negative symptoms in SZ (Kim et al., 2020; Murphy et al., 2020). However, these positive association studies have used different scales for clinical assessment (PANSS) than ours (SAPS & SANS). In fact, neuroimaging evidence suggests that MMN deficit in SZ is non-linear but progressive (Salisbury et al., 2007) which results in significantly and consistently deficient MMN amplitude in SZ but independent of severity of psychopathology and illness duration (Erickson et al., 2016). So, it is likely for SZ patients to have MMN deficits which may or may not correlate with psychopathology over the illness course.

For the unaffected FDR, schizotypy score, especially the cognitive-perceptual factor of schizotypy—that includes made up Ideas of Reference, Magical Thinking, Unusual Perceptual Experiences, and Paranoid Ideation—negatively correlated with MMNd and MMNf amplitudes. This finding is interesting because very few studies have examined MMN in SZ siblings/first-degree relatives with schizotypy. Our finding reiterates previous observations of reduced MMNd amplitude with higher eccentric perception (schizotypy dimension) in non-clinical participants (Donaldson et al., 2021). However, unlike us, this study did not find a correlation between MMNf amplitude and schizotypy. Another study reported reduction in MMNf amplitude in individuals with schizotypal personality disorder without finding any correlation between severity of schizotypy and degree of suppression of MMNf amplitude (Niznikiewicz et al., 2009), indicating that MMNf amplitude is potentially affected by schizotypal traits. However, contrary to expectation, a multi-feature (duration, pitch/frequency, intensity deviants) MMN study on healthy participants who undertook a schizotypal personality questionnaire (brief version), reported a positive trend-level correlation between MMNf amplitude and suspiciousness trait of schizotypy (Broyd et al., 2016). From the present study and existing findings in the literature, it is clear that MMN and schizotypy are potentially linked and deserve a nuanced examination. Schizotypy in non-clinical populations of healthy controls and unaffected FDRs may differ in their relation to MMN deviant types (duration, frequency, intensity, etc.). Furthermore, schizotypy is not a unitary construct; positive (odd, eccentric, paranoid traits) and negative (social disconnection, isolation, disinterest, etc.) schizotypy may have differential effect on the MMN deviant types.

This study has several limitations. Our study did not have a follow up component; especially for (a) SZ, noting relapse, and (b) unaffected FDR, noting transition or non-transition to psychosis, would have yeilded precious insight into stability and potential worsening of MMN with change in symptoms. Recording data from more number of channels would have enabled us to do advanced analysis examing how and whether cortical generators of MMN vary across SZ, HC and unaffected FDR, and whether this activity varies between chronic/medicated (cSZ) and early course/nonmedicated (dSZ) samples. For correlational analysis, we chose a one-tailed test to check our directional hypothesis in line with several recent reports (Madelung et al., 2022; Patil et al., 2017; Sugimura et al., 2021), though doing so renders the findings statistically weak. Given the versatile study sample of drug-naïve/drug-free and chronic-medicated patients, and the unaffected FDR participants with schizotypy scores, our goal was to test for possibility of clinically meaningful correlations that can indicate to future investigations with large samples whether to explore role of schizotypy and illness chronicity—and not just symptom severity— with respect to MMN.

Our study acknowledges duration deviant MMN as a more robust measure of MMN deficit in schizophrenia than the frequency deviant MMN. Duration deviant MMN correlated with illness chronicity, and we also note MMNd amplitude could be more impaired in hallucinating SZ patients. MMN latency neither significantly differs among groups nor does it show any meaningful correlation with any of the clinical features in SZ. This could explain why very few studies examine and report latencies. Unaffected FDR have MMN comparable to healthy controls. However, it is interesting to note that unaffected FDR with schizotypal personality traits (of made up of ideas of reference, magical thinking, unusual perceptual experiences, and paranoid ideation) have worse duration and frequency MMN amplitude compared to their counterparts who don’t express schizotypy. As schizotypy (with positive and negative dimensions) and MMN (with several deviant types) are nuanced subject matter, future studies should contextually examine how and whether these interact in HC and unaffected FDR.

## Data Availability

Data from this study are not publicly available due to ethical and privacy restrictions. However, data will be made available upon reasonable request from the corresponding authors.

## CREDIT AUTHOR STATEMENT

Author GVS designed the study. Authors AB, VSK, SMA & VSS collected the clinical data. Clinical symptom ratings were done by AB, SMA & VSK. Authors GV & JCN supervised the clinical assessment and ascertained the clinical ratings. ERP experiment was designed and implemented by AB and HN. ERP data was acquired by AB, HN and SMA. DK supervised the ERP experiment and ERP data analysis. AB analysed the ERP data and performed the statistical analyses. Authors AB & GVS managed the literature search and wrote the first draft of manuscript. All authors revised and optimized further versions of the manuscript. All the authors have contributed to and have approved the final manuscript.

## ACKNOWLEDGEMENTS

Anushree Bose is supported by Department of Biotechnology (DBT)-Wellcome Trust India Alliance Early Career Fellowship grant (IA/CPHE/19/1/504591). Venkataram Shivakumar acknowledges the support of Department of Biotechnology (DBT) - Wellcome Trust India Alliance Early Career Fellowship grant (IA/CPHE/18/1/503956). Vanteemar S Sreeraj acknowledges the support of the India-Korea joint program cooperation of science and technology by the National Research Foundation (NRF) Korea (2020K1A3A1A68093469), the Ministry of Science and ICT (MSIT) Korea, and the Department of Biotechnology (India) (DBT/IC-12031(22)-ICD-DBT). Ganesan Venkatasubramanian acknowledges the support of Department of Biotechnology (DBT) - Wellcome Trust India Alliance grants (500236/Z/11/Z and IA/CRC/19/1/610005).

