## Supplementary Table 1 for "Duration and Frequency Mismatch Negativity in Schizophrenia, unaffected first-degree relatives, and healthy controls"

**Supplementary Table 1: Demographic and clinical variables of cSZ (n=29) and dSZ (n=23) groups**

|  | Chronic SZ (cSZ) (n=29) | Drug naïve/free FZ (dSZ) (n=23) | Statistics | Uncorr. p |
| --- | --- | --- | --- | --- |
| Age (years) | 32.03±10.56 | 33.09±6.85 | U=227 | 0.297 |
| Sex | M=17;F=12 | M=11;F=12 | X**^2^**(1)=0.44 | 0.57 |
| Years of Education (years) | 13.55±3.79 | 10.52±4.72 | U=201 | 0.014* |
| Duration of Untreated Psychosis (months) | 16.07±25.37 | 37.17±30.13 | U=164 | 0.002* |
| Total Duration of Illness (months) | 109.86±74.47 | 44.70±33.94 | U=143 | <0.001* |
| Total SAPS | 33.79±11.83 | 24.65±7.82 | t=3.19 | 0.002* |
| Total SANS | 24.45±17.63 | 24.74±18.80 | t=0.50 | 0.95 |
| Hallucination Score (SAPS) | 16.28±4.60 | 11.48±3.67 | t=4.07 | <0.001* |
| Olanzapine Equivalent (n=29) | 24.36±17.02 | – | – | – |

* – Statistically significant at 0.05, uncorrected p; X**^2^** – Chi-square; U – Mann Whitney U test; t – Independent sample t-test
